# Autoimmunity in Satoyoshi Disease: a Systematic Review

**DOI:** 10.1101/2022.07.05.22277272

**Authors:** Vinícius Viana Abreu Montanaro, Julián Solís-García del Pozo, Thiago Falcão Hora, Beatriz Helena León, Carlos de Cabo, Javier Solera

**Affiliations:** SARAH Network of Rehabilitation Hospitals, Brasilia, Brasil. Programa de PósGraduação em Neurologia, Universidade Federal Fluminense, Niterói Brazil. Radboud University, Nijmegen, The Netherlands; Department of Internal Medicine, Unit of Infectious Diseases. Complejo Hospitalario Universitario de Albacete, Albacete, Spain; SARAH Network of Rehabilitation Hospitals, Brasilia, Brasil; Department of Pediatrics and Rheumatology, Universidad San Francisco de Quito, Quito, Ecuador; Research Department, Neuropsychopharmacology Unit, Complejo Hospitalario Universitario de Albacete, Albacete, Spain; Department of Internal Medicine, Complejo Hospitalario Universitario de Albacete, Albacete, Spain. Department of Medical Sciences, Faculty of Medicine, Universidad de Castilla – La Mancha, Albacete, Spain

## Abstract

**Background:** Satoyoshi syndrome (SS) is a rare multisystem disease. Although its cause is unknown, an autoimmune etiology has been postulated. In this paper we carried out a systematic review of all published cases of SS to evaluate the available evidence to support that autoimmune hypothesis.

**Methods:** We carried out a systematic review of the published cases of SS following the recommendations of the PRISMA statement for systematic reviews. We searched for SS cases in PubMed, the Web of Knowledge (WOS) and Scopus up to January 2022, using keywords “Satoyoshi syndrome” or “Komuragaeri disease”. Data on symptoms, associated autoimmune diseases, presence of autoantibodies and response to treatment were collected.

**Results:** 77 patients from 57 articles published between 1967 and 2021 were included. 59 patients were women. The mean age at diagnosis was 21.2 years. All cases had painful muscular spasms and alopecia. Other frequent manifestations included: diarrhea, malabsorption, growth retardation, amenorrhea and bone deformity. SS was associated with other autoimmune diseases: myasthenia gravis (2 patients), autoimmune thyroiditis (one patient), idiopathic thrombocytopenic purpura (one patient), atopic dermatitis (one patient), bronchial asthma (one patient) and lupus erythematosus (one patient). Autoantibody determinations were performed in 39 patients, of which 27 had positive results. The most frequently detected autoantibodies were antinuclear antibodies (21 patients). Other less frequently found auto-antibodies were: anti-acetylcholine receptor antibodies (7 patients), anti-DNA antibodies (5 patients), antithyroid antibodies (3 patients), anti-GAD (2 patients) and anti-gliadin antibodies (2 patients). Pharmacological treatment was reported in 50 patients. Most of them improved with corticosteroids (33 patients), immunosuppressants (9 patients) and immunoglobulins (10 patients), or a combination of these medications.

**Conclusions:** SS is associated with other autoimmune diseases and a variety of autoantibodies. Improvement after corticosteroid or other immunosupressant treatment was observed in 90% of cases. These data support an autoimmune etiology for SS. More studies are necessary, including the systematic determination of autoantibodies in all patients with SS to help us advance in our understanding of this disease.

## 1. Introduction

Satoyoshi syndrome (SS) is a rare multisystem disease of unknown etiology, characterized by painful muscle spasms, diarrhea, endocrinopathy, alopecia, and skeletal abnormalities [1,2]. Most cases were described in Asian children, and almost all were sporadic. Since the original description in 1967, few cases have been reported [3]. A clinical course without treatment may result in severe disability or death [1]. Despite this, the underlying pathophysiology of the disease and the clinical course are not fully understood.

Since the description of the syndrome, therapeutic strategies include muscle relaxants and antiepileptics to relieve muscular manifestations that can be very troublesome and cause significant disability. However, this type of treatment cannot relieve other disease manifestations, such as digestive symptoms or alopecia. Corticosteroids or other immunotherapies have been found to improve muscular manifestations as well as other pathological expressions of the disease. [2].

In addition to the immunosuppressive treatment, in recent years, this disease has been associated with serum autoantibodies known to other diseases, such as antinuclear antibodies [4,5]. Histopathological examination of the lesions detected in SS patients has also revealed signs of tissue inflammation. All the above mentioned findings suggest an autoimmune pathogenesis to this rare disease [3,6,7].

The objective of the present work is to offer an overview of all the available evidence pointing to an autoimmune origin in the pathogenesis of Satoyoshi syndrome.

## 2. Methods

We carried out a systematic review of the published cases of SS following the recommendations of the PRISMA statement for systematic reviews [8].

### 2.1. Search strategy

We reviewed all papers included in PubMed, Web of Knowledge (WOS), and Scopus until January 2022 using the keywords “Satoyoshi syndrome” or “Komuragaeri disease”, with no restrictions as to year of publication or language. The references from the articles found by these searches were reviewed to identify additional records. Papers published in languages other than English were translated using an automated program (Google translator). The literature search and inclusion of case reports was conducted by two authors (VM and JSGP) independently. In case of disagreement, the authors reached the final decision by consensus among all authors. The cases published in more than one article were considered a single case.

### 2.2. Inclusion of studies

Inclusion criteria were:

- Case records of Satoyoshi syndrome were defined by a compatible clinical picture consisting mainly of muscle spasms and alopecia (after having ruled out other possible causes), as well as some other manifestations such as:
  - Digestive symptoms in the form of diarrhea, carbohydrate malabsorption, or abdominal pain not related to muscle spasms.
  - Amenorrhea
  - Weight loss or growth retardation in the case of children
  - Skeletal alterations

Exclusion criteria were the following:

- Absence of individualized clinical case information necessary to confirm a diagnosis of SS, as it occurs in case series with aggregated data.

### 2.3. Data extraction

All papers were independently analyzed by two authors and the data were cross-compared at the end, with any discordance settled after debate.

The data extracted from each case were:

- Year of publication
– Epidemiological characteristics: age at onset of symptoms, age at diagnosis of SS, sex, country of origin, and ethnicity.
- Clinical data: a) Signs and symptoms including alopecia, muscle manifestations, diarrhea, or other digestive symptoms, growth retardation, skeletal alterations, amenorrhea, and anemia, b) Presence of other immune-associated diseases.
- Presence of autoantibodies (antinuclear antibodies, anti-DNA, anti-GAD, anti-acetylcholine receptor, antithyroid, SSA, SSB as well as other types of autoantibodies), results of biopsies and other immunological tests. The information was collected a*s* a positive or negative result, or not reported.
- Treatments received, including muscle relaxants, antiepileptic drugs, corticosteroids, other immunosuppressants, immunoglobulin therapy, or a combination of these drugs.
- Duration of treatment.
- Outcome: Mortality (death was considered SS-related if unattributable to another cause).

### 2.4. Statistical analysis

Statistical analyses were performed using the SPSS IBM version 21 program (IBM Corp., Armonk, NY). Categorical data were presented as frequencies and numerical variables with non-normal distribution as median and interquartile ranges. Correlations between variables were performed using the Chi-square test for categorical data and the Mann Whitney U-test for numerical data. Significance was set at p< 0.05.

## 3. Results

The searches in MEDLINE, Scopus, and WOS yielded 59, 68, and 79 articles, respectively. Sixteen additional works were retrieved by reviewing the bibliographies from the previous articles. A total of 77 cases of Satoyoshi syndrome were identified from 57 records [3-7, 9-60] and 2 personal communications. 14 records provided additional information on cases already reported in other references [61-74]. 13 were records not found [75-87] (Fig. 1, flow diagram). Of those 77 patients, 59 were women (76.6%). Median age at diagnosis was 17 years (15.5 years for women and 23for men, p=0.042). Median age at beginning of symptoms was 10.5 years (10 years for women and 13.5years for men, p=0.003).

**Figure 1:**
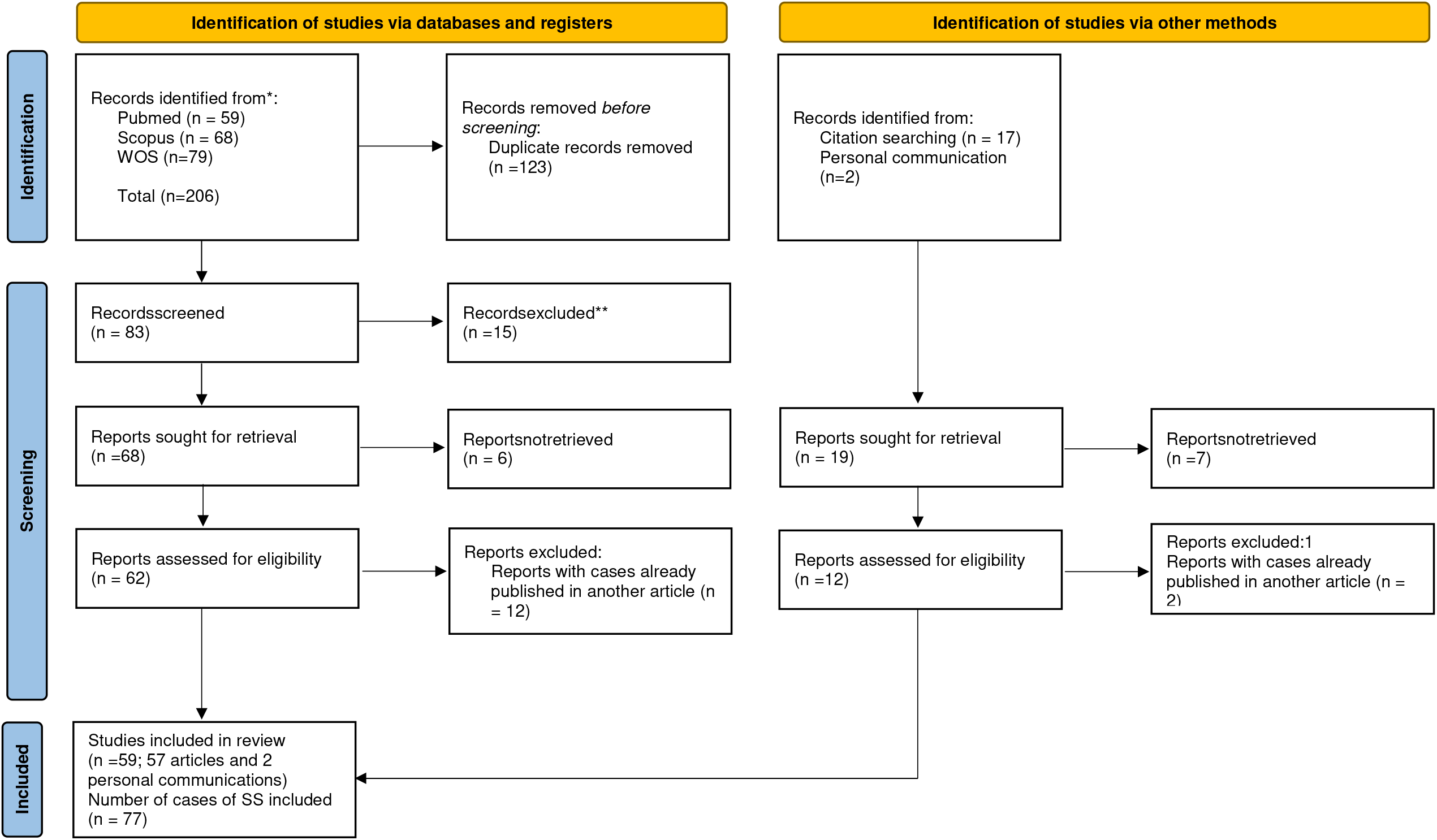
PRISMA 2020 flowdiagram [8].

### 3.1. Clinical features supporting the autoimmunity hypothesis

SS predominates in women, with a female to male ratio of approximately 3.3:1. The age of onset was under 20 years in 83% of cases. Twenty-nine cases were Japanese patients (37.7%), and 11 cases were from China (14.3%). Overall, 55.8% were patients of East Asian ethnicity (Japanese, Thai, Chinese, Filipino, or Asian-American).

Muscular manifestations were present in 100% of patients. They usually consisted of painful muscle cramps or spasms affecting limbs, trunk, and even masticatory muscles [46]. Satoyoshi (1978) suggested involvement of abnormal discharges from the neurons of the anterior medullary horn in muscular symptoms [3]. Electrophysiological studies point to deregulation at the alpha motor neuron level, leading to involuntary muscle contractions [9].The differential diagnosis of these manifestations includes stiff-person syndrome and Isaac’s syndrome. Both entities are associated with autoimmune phenomena. Stiff-person syndrome is linked to the presence of anti-GAD antibodies also reported in two cases of SS [11,65]. Unlike these two diseases, in SS, muscle contractions are painful, and other manifestations such as myokymias or fasciculations are absent [10]. Other findings also suggest an immune-based pathogenesis for muscle symptoms. Aghoram reported reactivity to unknown neuronal antigens, monkey cerebellum, and peripheral nerve tissue [12]. In other SS cases, striate muscle antibodies were found [13, 14]. Additionally, in at least two cases, antibodies directed against brain lysates were detected [15,66]. In one patient also diagnosed with myasthenia, an inflammatory infiltrate was seen in muscle biopsies [16].

The second manifestation that invariably appears in patients with Satoyoshi syndrome is alopecia, which in many cases affects not only the head, but also the eyebrows, eyelashes, and body hair (alopecia universalis). The principal differential diagnosis in the alopecia of SS patients is alopecia areata, an autoimmune disease. Skin biopsies inSS patients have detected inflammatory infiltrate indistinguishable from alopecia areata [7, 13, 17, 18, 19, 20].

The third cardinal manifestation of SS is digestive symptoms, mainly in the form of diarrhea with malabsorption. These manifestations also appear in other entities with an altered immune or inflammatory response, such as celiac disease, inflammatory bowel disease, and autoimmune enteritis [88]. In cases of SS displaying digestive symptoms, gastrointestinal biopsies show the presence of a predominantly lymphocytic inflammatory infiltrate [6, 15, 21, 22]. Antigliadin antibodies have also been detected [13].

Other clinical manifestations include bone deformities, growth retardation, and amenorrhea. Bone deformities are a result of muscular contractions, mainly in growing individuals with non-closed bony metaphyses [6, 40]. Growth retardation in children and weight loss in adults could be explained by diarrhea, malabsorption, and increased energy expenditure due to recurrent muscle contractions. Amenorrhea was present in 27 women (45.8%). 10 out of 13 cases (76.9%) showed positive autoantibodies [5, 11, 19, 22 – 24,26 – 29]. Delayed puberty could be explained by growth retardation and low weight, although uterine hypoplasia has also been described [3, 16].

SS has been observed to co-exist with other immune-based diseases in the same patient, such as autoimmune thyroiditis [25], myasthenia gravis [16, 26], idiopathic thrombocytopenic purpura [27], atopic dermatitis [28], and bronchial asthma [28]. Solera et al. reported the possible association of one particular gene with the immune response in a SS patient [64].

### 3.2. Autoantibody detection and other analytical data

Table 1 and figure 2 summarize the antibodies found in patients with Satoyoshi syndrome. In 39 of the 77 cases, the presence of autoantibodies was investigated. 27 showed some positive serum antibodies (69.2%), and 12 did not. Of these 27 antibody-positive patients, 14 were 1-antibody positive, 11 were 2-antibody positive, and two were 3-antibody positive. There were no differences in sex (25.6% versus 21.1% were male; p=0.634) nor median age of onset (12 vs. 10; p=0.131) between patients with autoantibody data available and those without. Most of the cases reporting autoantibody data were from recent years.

**Table 1:** Autoantibodies detected in patients with SS

| **Autoantibodies** | **No data reported** | **Negative** | **Positive** | **% of reported positives** |
| --- | --- | --- | --- | --- |
| Antinuclear antibodies | 41 | 15 | 21 | 58.3% |
| Anti-DNA | 61 | 11 | 5 | 31.25% |
| Antithyroid antibodies | 65 | 9 | 3 | 25% |
| Acetylcholine receptor antibodies | 60 | 10 | 7* | 41.18% |
| Anti GAD | 64 | 11 | 2 | 15.38% |
| SSB | 70 | 6 | 1 | 14.29% |
| Antigliadin | 72 | 3 | 2* | 40% |

\* Antigliadin antibodies were found for the patient reported by Solera et al. [63] (unpublished data). Anti-acetylcholine receptor antibodies were found for the patient reported by Merino de Paz [13] (unpublished data).

**Figure 2:**
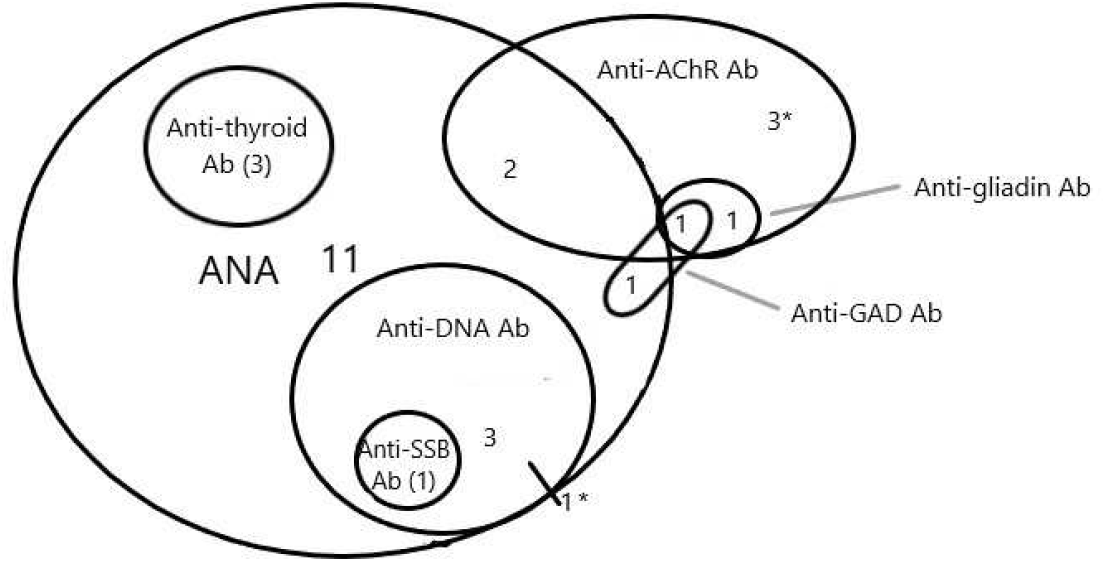
A graphic representation of the coexisting autoantibodies in patients with SS. *For one case with anti-DNA Ab and another case with anti-AChR Ab, results regarding the presence of ANA were unavailable

**Table 2:** Summary of the clinical features supporting the autoimmune origin of Satoyoshi syndrome

| **Clinical features** | **Data supporting the hypothesis that SS is an immune/inflammatory disease** |
| --- | --- |
| Sociodemographic characteristics | – Female to male ratio of approximately 3.3:1.<br>– The most common age of onset is between 10 and 20 years. |
| Muscle manifestations | – Anti-GAD antibodies [65, 11] in two cases.<br>– Similar to other immune-mediated diseases (Stiff-man syndrome and Isaac's syndrome).<br>– Reactivity to unknown neuronal antigens in monkey cerebellum and peripheral nerve tissue. Anti-striated or striated muscle antibodies in three cases [11, 13, 14].<br>– Antibodies against brain lysates in two cases [15,66]. |
| Alopecia | – Biopsies with findings indistinguishable from alopecia areata [7, 13,17, 18,19, 20] |
| Diarrhea | – Diarrhea with malabsorption is similar to that of other diseases such as celiac disease. Gastrointestinal biopsies with a predominantly lymphocytic inflammatory infiltrate [21, 6, 15, 22].<br>– Antigliadin antibodies were detected in two cases [13, 63*] |
| Association with other autoimmune-based diseases | – Association with other immune-based diseases in the same patient: autoimmune thyroiditis [25], myasthenia gravis [16, 26], idiopathic thrombocytopenic purpura [27], atopic dermatitis [28], bronchial asthma [28].<br>– Association of one particular gene with the immune response in one patient. |
| Response totreatment | – Muscle relaxants and antiepileptic drugs do not improve all symptoms.<br>– Corticosteroids and immunosuppressive treatment can improve muscle symptoms, diarrhea, and alopecia.<br>– Only two of the eight patients who died [3,22,35,56] received corticosteroid treatment. These two patients died due to neuroleptic malignant syndrome attributed to midazolam [35] and intestinal perforation [56]. |

\* Antigliadin antibodies were found for the patient reported by Solera et al. [63] (unpublished data).

Within the patients with autoantibody data available, there were differences in age of onset (median 13 versus 8; p= 0.026) between those with some positive autoantibody and those without. Regarding the percentage of patients treated with corticosteroids, there were no differences between the group of antibody-positive patients and the non-positive group (p=0.642). However, immunoglobulins (39.1% vs. 12.5%; p= 0.222) and other immunosuppressants (34.8% vs. 0%; p= 0.076) were used more frequently among those with some autoantibody positivity (without reaching statistical significance). The rate of patients in whom dantrolene (13% vs. 25%; p=0.583) or antiepileptic drugs (18.2% vs. 37.5%; p=0.345) were used was higher among those without a positive autoantibody, although statistical significance was not found in any of these cases.

Antinuclear antibodies were positive in 21 patients, with antibody-titers ranging from 1/40 to 1/1280. 10 of these patients (45.5%) were also positive for some other autoantibody and1 of these was positive for three antibodies. The concomitant positive antibodies with ANA were: four anti-DNA, three antithyroid, two anti-acetylcholine receptor, one anti-GAD, and one SSB antibodies (Figure 2). Among the ANA-negative patients, four had anti-acetylcholine receptor, two had antigliadin, and one had anti-GAD antibodies. However, there were no significant differences in the treatments used for patients with positive ANA vs negative ANA antibodies.

### 3.3. Data on treatment and clinical evolution

We have data on pharmacological treatments administered to 50 patients (64.9%):33 received corticosteroids (66%), 9 others immunosuppressants (18%), 10immunoglobulins (20%), 11 dantrolene (22%), and 15 antiepileptics (30%). Symptomatic treatment with medications such as dantrolene, carbamazepine or botulinum toxin, were the mainstay of the first published cases [3], with mixed results. In most cases, muscular symptoms improved with corticosteroids, and the patient was able to lead a normal life with little interference from symptoms [6, 18, 29, 30 - 33]. Although full hair recovery was rare, some hair regrowth was reported in most cases [6, 34, 29, 30, 31, 32, 18, 33]. Digestive symptoms also responded to treatment with steroids, with the disappearance of diarrhea [6, 21, 31], and menstruation also reappeared in many patients [30, 31, 19, 33].

In the literature search, we found eight SS patients who died (10%) [3, 22, 35, 56]. Five out of these eight cases were described by Satoyoshi in 1978 [3]. The three other cases were described by Nagahama et al. [22], Adachi et al. [35] and Yuhong et al. [56]. Only two of the eight patients who died [35, 56] received corticosteroid treatment. The death of these two patients was due to neuroleptic malignant syndrome [35] and intestinal perforation [56].

## 4. Discussion

Autoimmune disorders are a very heterogeneous group of diseases which share the presence of antibodies reacting against autoantigens as their common denominator. In our work, we analyzed all the evidence supporting SS as a possible autoimmune disease.

Ethnic and geographic differences in the incidence of specific autoimmune diseases have been documented [89]. Consistent with this notion, SS seems to be more frequent in people of Asian origin. As occurs in most autoimmune diseases, SS is more frequent in women [90, 91].

The three cardinal manifestations of the disease (muscular spasms, alopecia, and diarrhea) are also present in other autoimmune disorders. In addition, mainly digestive and skin biopsies invariably show a predominantly lymphocytic inflammatory infiltrate without an obvious infectious cause.

The presence of autoantibodies is key for the diagnosis of autoimmune diseases. Of the 39 cases with autoantibody determinations, at least one autoantibody was detected in 27 patients (69.2%). The most frequent autoantibodies were the antinuclear antibodies: 53.8% of patients with at least one antibody detected and 58.3% of patients with ANA measurements. Markers of serological autoimmunity, such as anti-nuclear antibodies (ANA), are found in approximately 21-25% of the general population [92,93,94]. However, in a study conducted on the Chinese population, the overall prevalence of ANA was only 5.92% and correlated positively with age [95]. In our study, more than 50% of the patients with some autoantibody data were positive for ANA, which suggests an autoimmune origin for SS.

It is noteworthy that of the 7 out of 17 cases that were tested for anti-acetylcholine receptor antibodies, 41% were positive. Of these 7, only one patient was diagnosed with myasthenia gravis [26]. This patient, besides high levels of anti-acetylcholine receptor antibodies, also showed deposition of anti-C3 and anti-C9 immunocomplexes at motor endplates in a muscle biopsy. Despite the frequent appearance of anti-acetylcholine receptor antibodies in SS patients, their role in the disease is not yet known.

Both Endo at al. and Matsuura et al. found antibodies against brain and gastrointestinal tissues in the serum of three patients with Satoyoshi [15,66]. An important next step would be to demonstrate a specific antigen as target of these auto-immune reactions, which might also allow the isolation of a disease-specific antibody. Solera et al.[64] have suggested a gene potentially related to the immune response in SS.

The response to immunosuppression is another solid argument that supports the role of immunity in the pathogenesis of SS. Treatment with corticosteroids and immunosuppressants has improved the prognosis of all the manifestations of SS [2].

Despite our efforts in the present study to gather all the available immunological information on SS, many questions remain unanswered. Autoantibodies specific for SS have not yet been found, nor have we been able to correlate antibodies levels with severity of disease. As is the case with other rare diseases, the very low prevalence of SS has been an obstacle to research. An international collaboration of all possible specialists would be necessary to register patients with SS and take serum samples. In the future, autoantibody determination in all SS patients could help us advance our understanding of this disease.

## Data Availability

All data produced in the present study are available upon reasonable request to the authors

## 5. Acknowledgment

We thank Alexandra L. Salewski MSc. for expert English review of the manuscript.

## List of abbreviations

Ab: antibodies
ANA: AntiNuclear Antibodies
AChR: acetylcholine receptor
GAD: glutamic acid decarboxylase
SSB: single-stranded DNA binding proteins
SSA: anti–Sjögren’s-syndrome-related antigen A

